# Target temperature management and post-extracorporeal cardiopulmonary resuscitation outcome: A post hoc analysis of the SAVE-J II Study

**DOI:** 10.1101/2023.06.15.23291462

**Authors:** Jun Kanda, Shinji Nakahara, Akihiko Inoue, Toru Hifumi, Tomoya Okazaki, Migaku Kikuchi, Shoji Yokobori, Yasufumi Miyake, Naoto Morimura, Tetsuya Sakamoto, Yasuhiro Kuroda

## Abstract

**Background:** The conflicting results of previous analyses about hypothermia management in patients with out-of-hospital cardiopulmonary arrest have hindered the establishment of a uniform standard temperature setting for temperature control. This study investigated and compared the clinical outcomes of hypothermic (target temperature: 32–34°C) and normothermic (35–36°C) management of out-of-hospital cardiac arrest (OHCA) patients, treated with extracorporeal cardiopulmonary resuscitation (ECPR).

**Methods:** This secondary analysis of the SAVE-J II study, a retrospective, multicenter, registry study involving 36 participating institutions in Japan, was undertaken, and ECPR patients with a suspected cardiac etiology were included in this cohort. The primary outcome was survival at hospital discharge. Favorable neurological outcomes (5-point Glasgow-Pittsburgh Cerebral Performance Categories 1–2) constituted the secondary outcome. Multivariable logistic analysis, which was adjusted for potential confounders, was performed for the primary and secondary outcomes.

**Results:** Of the 949 participants of this study, 57% underwent hypothermic management. A total favorable neurological outcome at hospital discharge was identified in 164 patients (17%), and the survival rate was 35%. In multivariable analysis, with the primary and secondary endpoints as each dependent variable, and gender, age, witness, bystander CPR, electrocardiogram, low flow time, and causative disease as categorical covariates, hypothermic management compared to normothermic management in OHCA patients treated with ECPR, was not significantly associated with a favorable neurological outcome (adjusted odds ratio (aOR): 1.22: 95% CI: 0.85–1.74), but was associated with survival (aOR: 1.74: 95% CI: 1.31–2.32).

**Conclusions:** Compared to normothermic management, hypothermic management of OHCA patients treated with ECPR was not significantly associated with a favorable neurological outcome, but was associated with survival at hospital discharge.

## Clinical Perspective

1. What is new?
  1. This study showed that hypothermia (32–34°C) management of body temperature after ECPR induction resulted in better survival.
  2. In the secondary analysis, the prognosis of hypothermia management in cardiogenicity other than acute coronary syndrome (arrhythmia, myopathy, myocarditis, and other cardiac causes) was particularly good.
2. What are the clinical implications?
  1. Hypothermic management after cardiopulmonary arrest should be implemented if ECMO ensures stable circulatory dynamics.

## Non-standard Abbreviations and Acronyms

ACS: acute coronary syndrome, CPC: cerebral performance categories, cOR: crude odds ratio, aOR: adjusted odds ratio, CPR: cardiopulmonary resuscitation, ECG: electrocardiogram, ECMO: extracorporeal cardiopulmonary resuscitation, OHCA: out-of-hospital cardiac arrest, PEA: pulseless electrical activity, VF: ventricular fibrillation, VT: ventricular tachycardia.

## Background

Extracorporeal cardiopulmonary resuscitation (ECPR), which combines extracorporeal membrane oxygenation (ECMO) with conventional cardiopulmonary resuscitation (CPR), is a more aggressive cardiopulmonary resuscitation technique in patients with cardiopulmonary arrest, that facilitates early resumption of cerebral blood flow.^1^ Sakamoto et al. reported that in out-of-hospital cardiac arrest (OHCA) patients with ventricular fibrillation (VF) / ventricular tachycardia (VT) on the initial ECG, a treatment bundle including ECPR, therapeutic hypothermia, and intra-aortic balloon pumping (IABP) was associated with improved neurological outcome at 1 and 6 months after OHCA compared to that by conventional CPR; thus, ECPR may improve neurological outcomes.^2^ However, the selection of patients for ECMO and its management have not been adequately studied, and each institution currently operates following institutional criteria.^3^

In contrast, in 2002, the Hypothermia After Cardiac Arrest Study Group and Bernard SA reported that hypothermia management at 32–34°C and 33°C, respectively, resulted in better outcomes than did conventional temperature management, in patients with out-of-hospital cardiopulmonary arrest with prolonged loss of consciousness after resumption of heartbeat; these studies constituted the basis for various guidelines worldwide.^4–6^ However, additional studies on hypothermia management, such as the Targeted Temperature Management Trial (TTM)-1 Study in 2013 and the TTM-2 Study in 2021, found no difference in outcome between normothermic management, which avoids hyperthermia and maintains a normal temperature, and hypothermic management, which targets temperatures of 32–34°C, and recommended that there is no need to use hypothermic management.^7, 8^ However, in the 2019 HYPERION trial of patients with coma, after resuscitation from cardiac arrest with nonshockable rhythm, the 33°C hypothermic management group had better 90-day neurological outcomes compared to that of the 37°C normothermia management group.^9^ In addition, two reports on temperature management using the Japanese Association for Acute Medicine Out-of-Hospital Cardiac Arrest (JAAM-OHCA) Registry reported three severity categories (mild, moderate, and severe), that were calculated from variables including the electrocardiogram (ECG), witnessed arrest, hypoperfusion time, blood gas (pH and lactate), and the Glasgow Coma Scale score and averred that hypothermic management was effective in the moderate, but not mild and severe, categories.^10^ However, another report of patients on ECMO from the JAAM-OHCA Registry, showed that hypothermic management was ineffective.^11^ Previous analyses by disease severity have been performed; however, no disease-specific analyses have been conducted.^10^ These conflicting results have hindered the establishment of a uniform standard temperature setting for temperature control.^12^

This study was conducted with an aim to compare the outcome of hypothermic and normothermic cardiopulmonary resuscitation management under ECMO; thus, contributing to the development of methods for the management of body temperature after a cardiac arrest.

## Methods

### Study design

This study involved a secondary analysis of data that were provided by the SAVE-J II Study Group – a retrospective, multicenter registry study with 36 participating centers in Japan, that was based on enrolment in the University Hospital Medical Information Network Clinical Trials Registry and the Japanese clinical trial registry (registration number: UMIN000036490).^13^

The SAVE-J II Study included adult patients (age ≥18 years) who were transported directly to the emergency department of a participating facility for treatment of out-of-hospital cardiac arrest (OHCA) and underwent ECPR in the resuscitation room before return of spontaneous circulation (ROSC) between January 1, 2013, and December 31, 2018. Undergoing ECMO in addition to conventional CPR was defined as ECPR. Patients who underwent ECMO after ROSC, patients transferred from other hospitals, those with exogenous diseases, acute aortic dissection/aneurysm other than cardiogenic causes, and primary brain injury were excluded. Cases in which the cause of cardiopulmonary arrest could not be determined to be cardiogenic at the time of the visit, but was presumed to be cardiogenic, were not excluded in order to consider the indication for ECPR. Moreover, patients who declined to participate by proxy, such as family members, were excluded. Of the 1,646 patients who were enrolled in the SAVE-J II Study, those who died before ICU admission and were ineligible for temperature control (165 patients), whose temperature control information was unknown (608 patients), and whose target body temperature was not in the target temperature range (32.0–36.5°C; n=7) were excluded. Furthermore, we excluded patients with unknown cerebral performance categories (CPC), age, sex, witnesses, bystander CPR, electrocardiogram (ECG), low flow time, and diagnosis (86 patients), which were the variables used in the analysis (Fig. 1).

**Figure 1.**
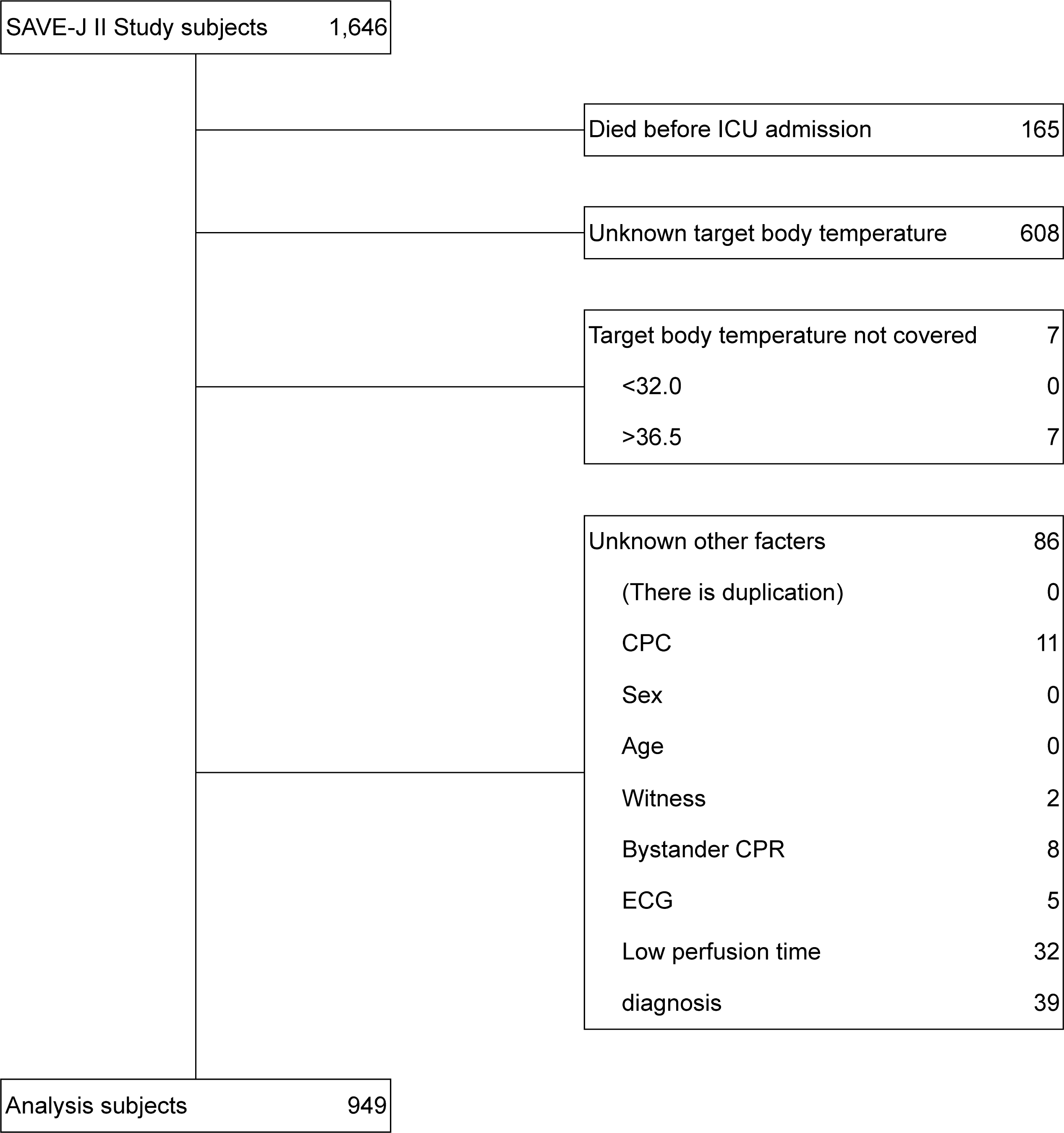
Flowchart depicting participant selection. ICU: intensive care unit, CPC: cerebral performance categories, CPR: cardiopulmonary resuscitation, ECG: electrocardiogram

### Data collection

Data were provided by the SAVE-J II Study and included information on age, sex, witnesses, bystander CPR, place of occurrence, ECG at scene and on arrival at the hospital, use of adrenaline and defibrillation, prehospital airway management, heart rate before starting ECMO, treatment-related factors, time course, cause of cardiac arrest, ROSC after hospital arrival, ECMO management information (which included temperature settings), duration of maintenance, and cooling method (use of heat exchangers). Details of the hospital and ICU length of stay, in-hospital mortality, and neurologic outcome (CPC) were also collected. The target temperature was defined as the ECMO set temperature; however, if the temperature setting was changed during ECMO management, the last set temperature was used as the target temperature. ROSC was defined as pulse confirmation that lasted for at least 1 minute. The estimated low-flow time was defined as the time from cardiac arrest to ECMO establishment, if the location of cardiac arrest was an ambulance, or as the time from calling an ambulance to ECMO establishment, if the location of cardiac arrest was a non-ambulance location. Causes of cardiac arrest were categorized as acute coronary syndrome, arrhythmia, myopathy, myocarditis, other cardiac causes, pulmonary embolism, and other non-cardiac causes.

### Endpoints

The primary endpoint was survival at 28 days. The secondary endpoint was the neurological outcome at 28 days, as assessed by the CPC (5-point Glasgow-Pittsburgh Cerebral Performance Categories): CPC1: able to work, CPC2: able to perform daily activities without assistance, CPC3: conscious but needs assistance in daily activities, CPC4: coma, and CPC5: death. In this study, CPC1–4 was defined as survival and CPC1–2 was defined as good neurological outcome for analysis.^14^

### Variables

The target body temperature during ECPR was the main independent study variable and was defined as the final temperature setting of ECMO (the temperature setting of ECMO is defined as the “target temperature” in 0.5℃ increments). The target body temperature was divided into 32.0–34.5°C (32–34°C group) and 35.0–36.5°C (35–36°C group) to enable an intergroup comparison. Besides this, each covariate was categorized as follows. Age was categorized into 64 years or younger and 65 years or older; ECG was categorized into 3 categories, VF and Pulseless VT, Asystole, and pulseless electrical activity (PEA); low flow time, defined as the time from the start of chest compressions to the point before ECMO induction, was categorized into 60 minutes or less and 61 minutes or more; the cause of cardiac arrest was categorized as acute coronary syndrome (ACS), cardiogenic except ACS (arrhythmia, myopathy, myocarditis, and other cardiac causes), and non-cardiogenic (pulmonary embolism and other non-cardiac causes). Patients with serious diseases other than exogenous and cardiogenic illnesses (such as acute aortic dissection/aneurysm, primary brain damage, etc.) were excluded from the SAVE-J II Study.

### Analysis

With the primary and secondary endpoints as each dependent variable and the target temperature, sex, age, witnessed arrest, bystander CPR, ECG, low flow time, and causative disease as categorical covariates, a multivariate logistic analysis was performed as the main analysis of this study to calculate the adjusted odd ratio (aOR) and 95% confidence intervals (CIs) for each variable. In addition, a model with an interaction term was created by assuming that the association between the target temperature and outcome was influenced by the ECG waveform and causative disease, and that there was an interaction.^4, 5, 7–9, 15^ If there was a significant interaction, further secondary analysis was performed after stratification by that factor.

### Sample size

As this study constitutes a multivariate logistic analysis with 7 factors as covariates in the main and secondary analyses, we determined that a sample size of at least 10 cases per factor, for a total of 70 cases, was necessary.

### Ethics

The SAVE-J II Study was approved by the institutional review board of Kagawa University (approval number: 2018-110). For all participating centers, the requirement for patient consent was waived because of the retrospective nature of the study. The study was approved by the Teikyo University School of Medicine Research Ethics Committee (approval number: 23-005).

## Results

### Description of participants

In this study, a total of 949 patients were included, which comprised 541 participants in the 32–34°C group and 408 participants in the 35–36°C group. The percentages of good neurological outcome and survival were 19% and 41%, respectively, for the 32–34°C and 35– 36°C groups, compared to 15% and 27% for the 35–36°C group.. (Table 1).

**Table 1.**
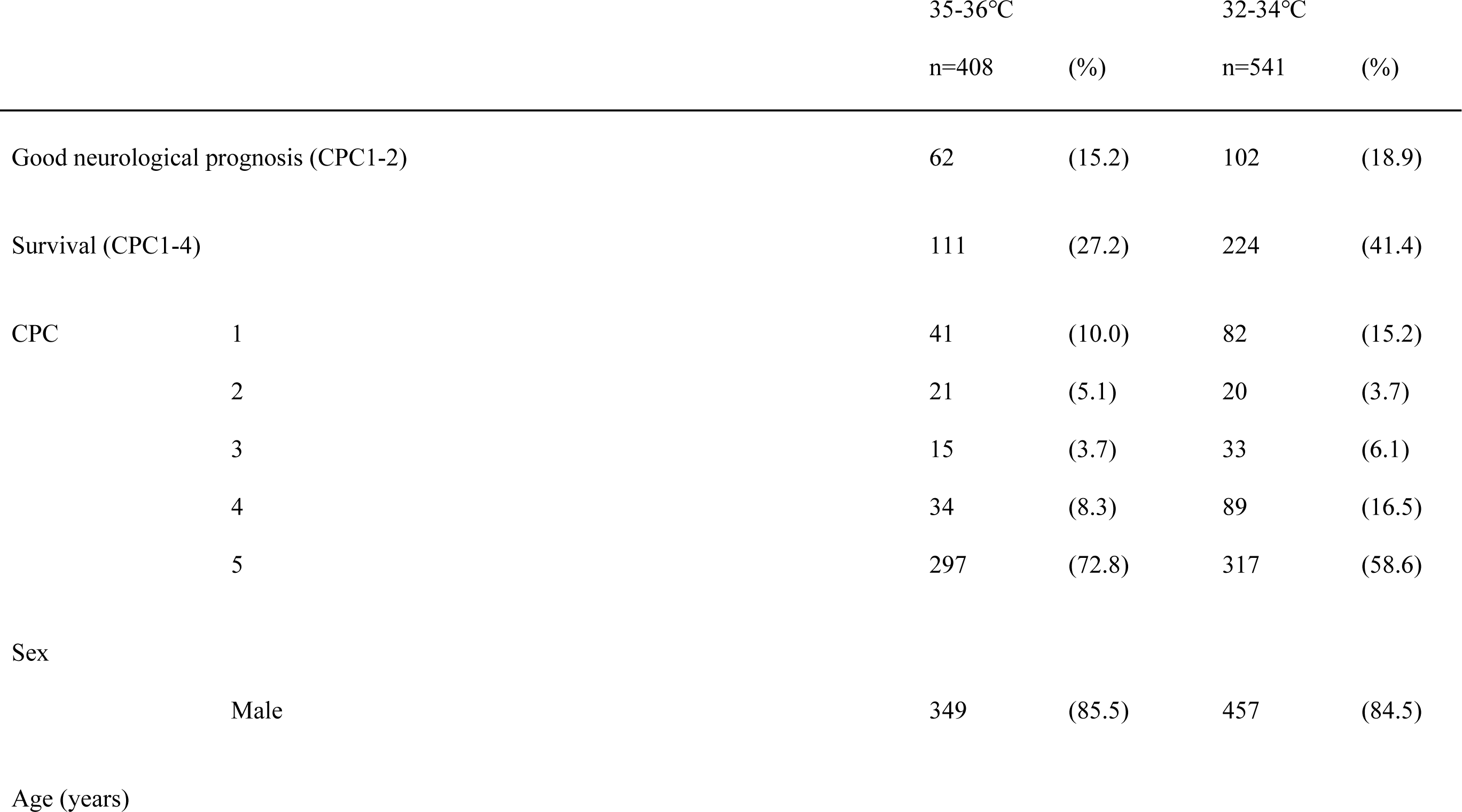
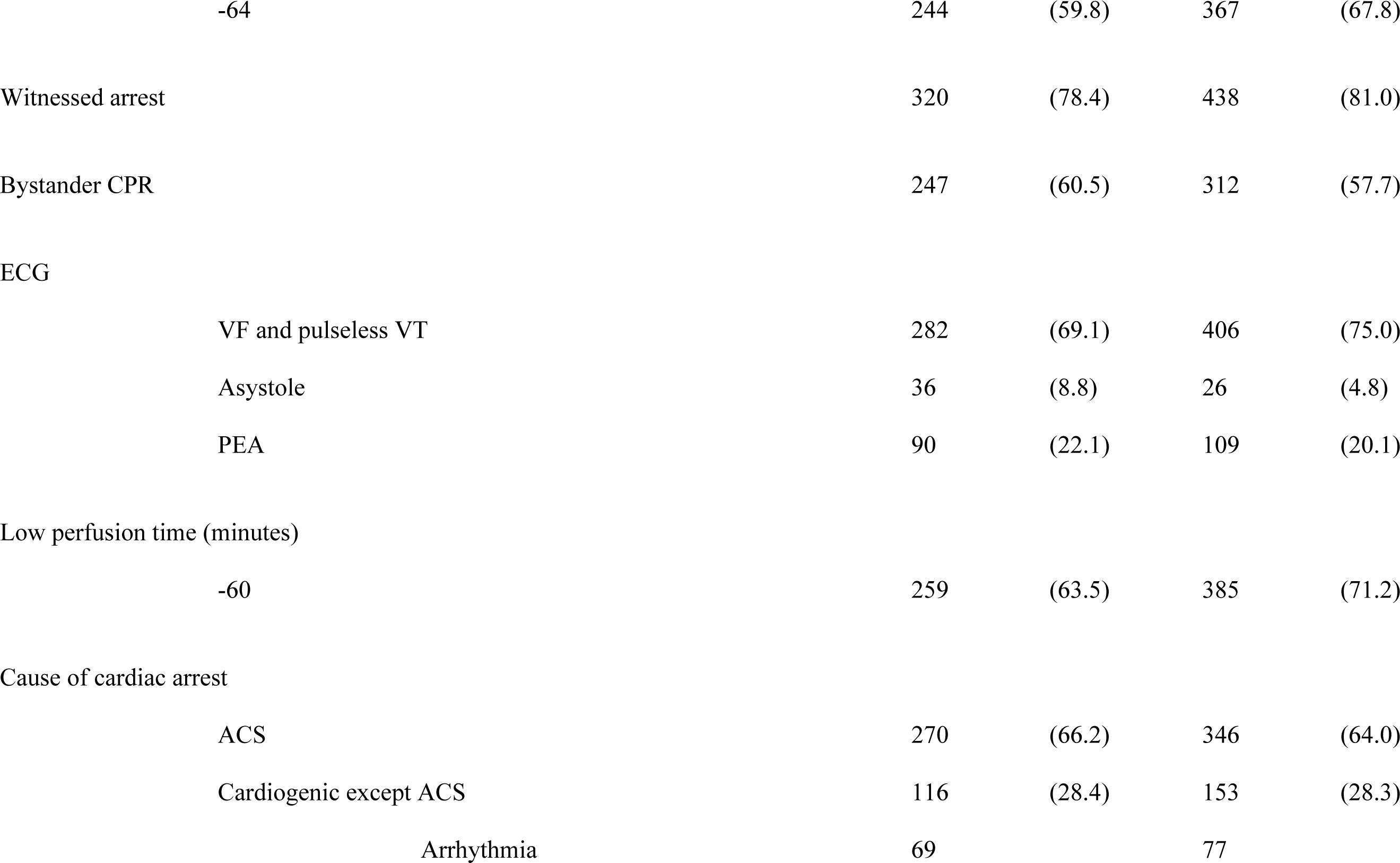
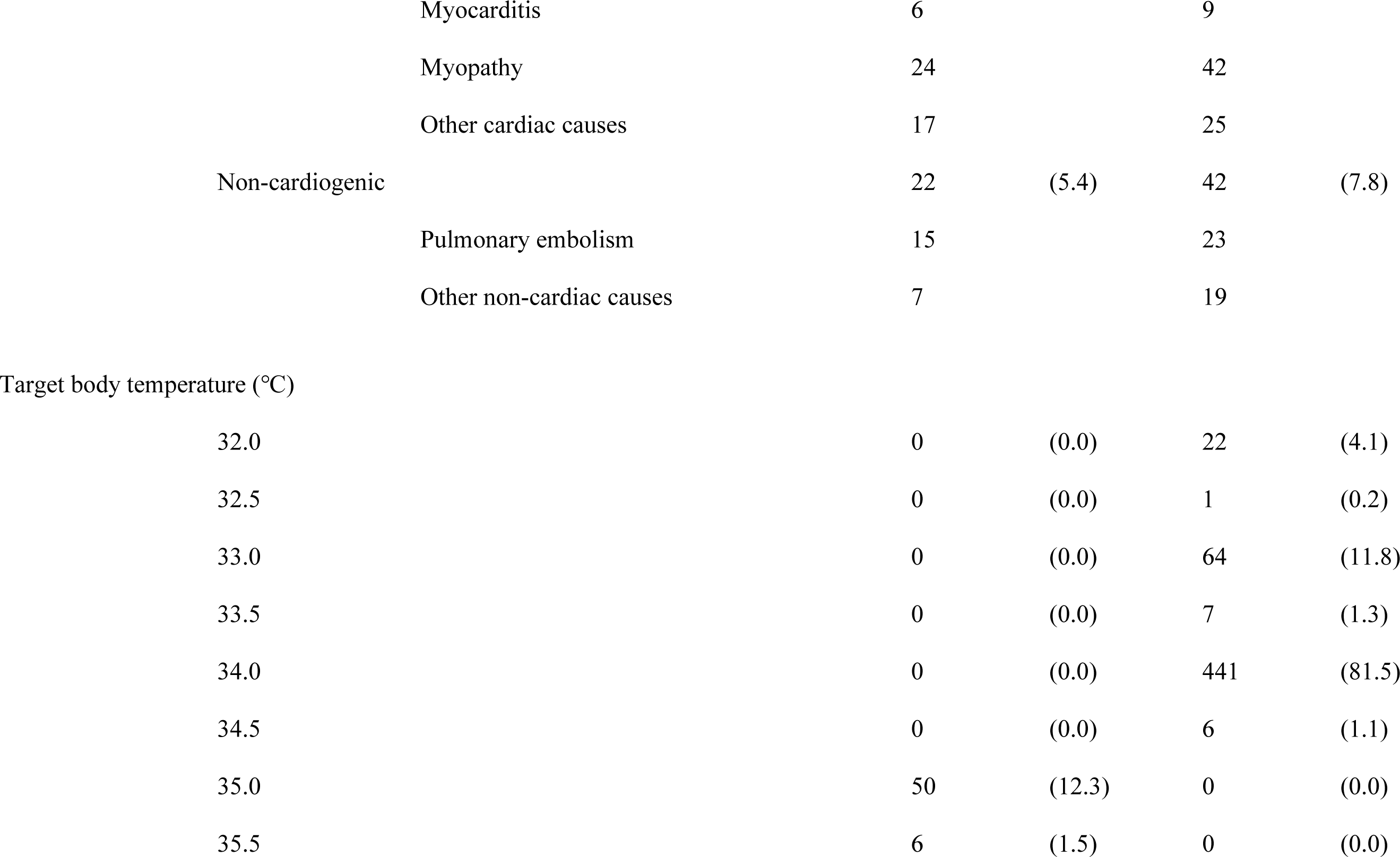
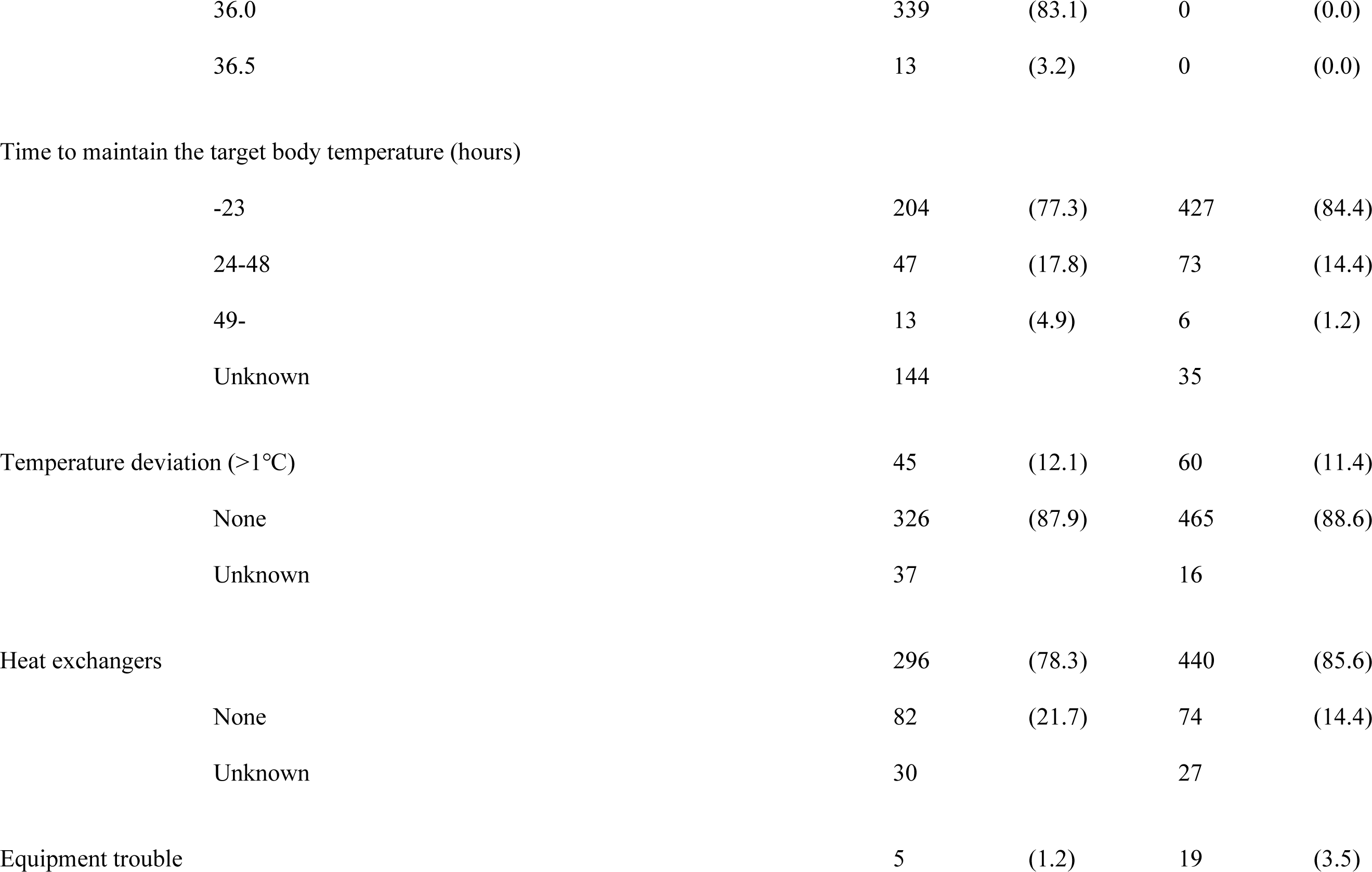
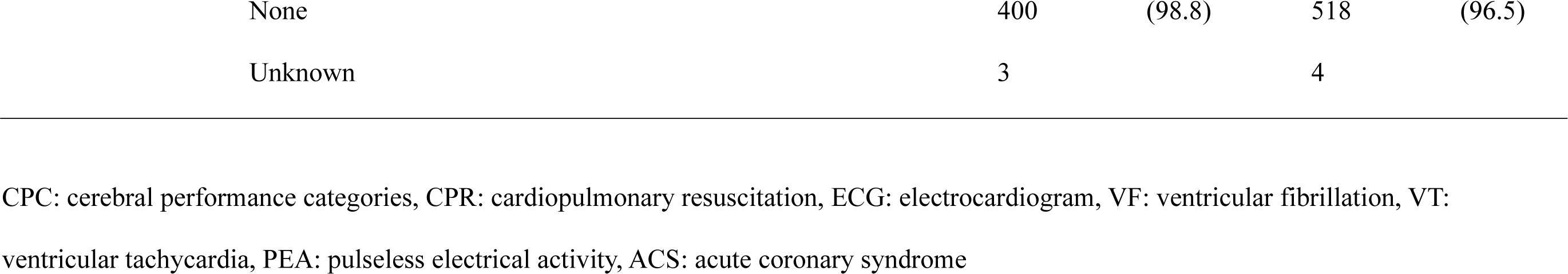
Description of the participants’ characteristics.

In both groups, 60–70% of participants were 64 years old or younger and approximately 85% were male. Moreover, there were 80% witnessed arrests and approximately 60% received bystander CPR. Analysis of the ECGs revealed that 70–75% showed VF and pulseless VT, which are shock-adapted. Target body temperatures of 34.0°C for 32–34°C and 36.0°C for 35–36°C were attained in greater than 80% of the cohort. In each group, heat exchangers were used in approximately 80% of cases, with similar deviations of more than 1℃ in approximately 10%, and a small number of equipment problems in less than 3% of cases. In both groups, ACS was the causative disease in approximately 65%, cardiogenic other than ACS in just under 30%, and noncardiogenic in 5–8% of cases. (Table 1)

### Main analysis

The aOR for good neurological outcome at 32–34°C versus 35–36°C at target body temperature was 1.2 (95% CI: 0.8–1.7), and the aOR for survival was 1.7 (95% CI: 1.3–2.3). Good neurological outcome was associated with female sex, bystander CPR, and ECG (VF and Pulseless VT). Survival was associated with target body temperature (32–34°C), witnessed arrest, ECG (VF and Pulseless VT), and low flow time (<60 minutes). (Table 2).

**Table 2.**
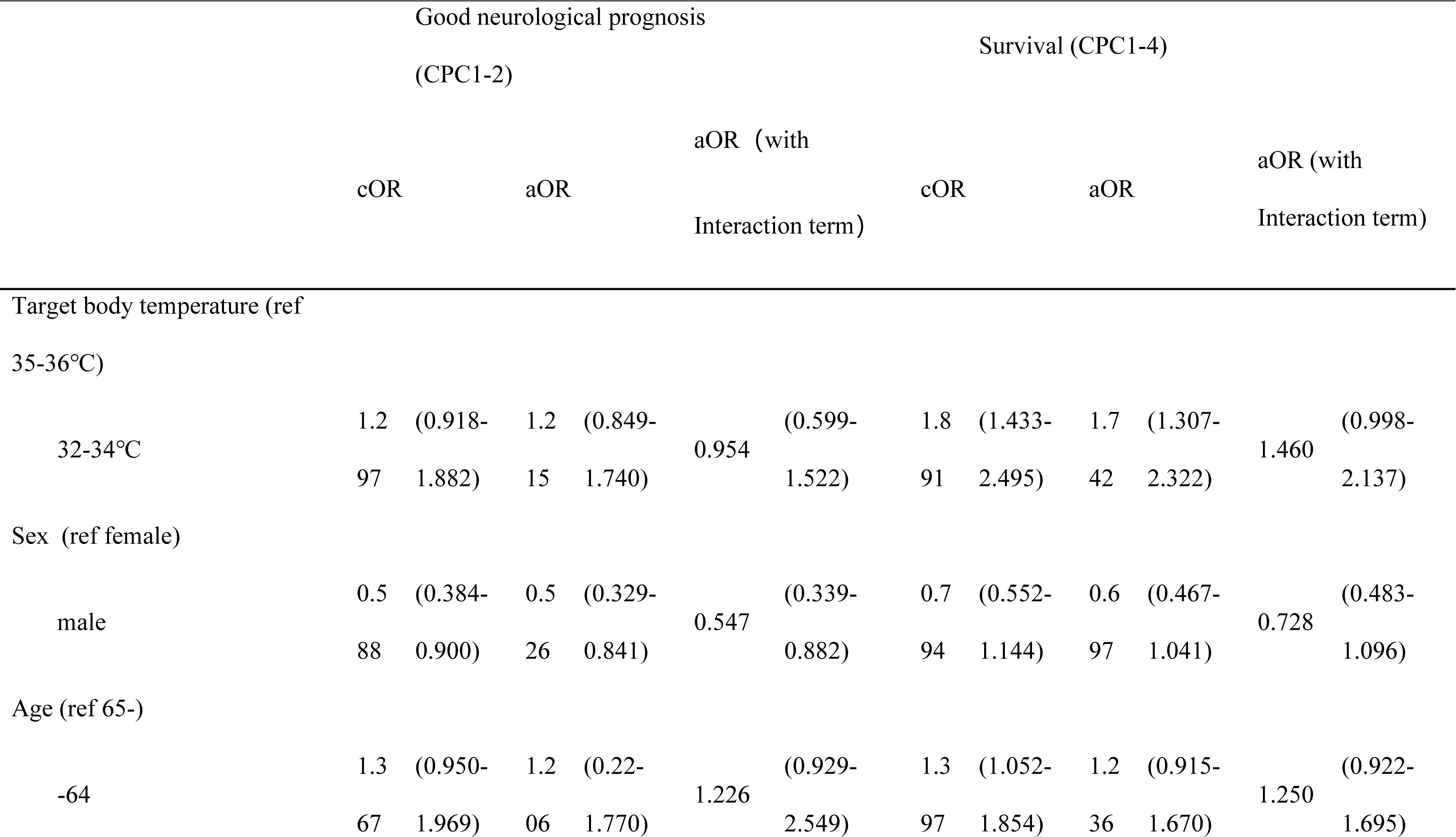
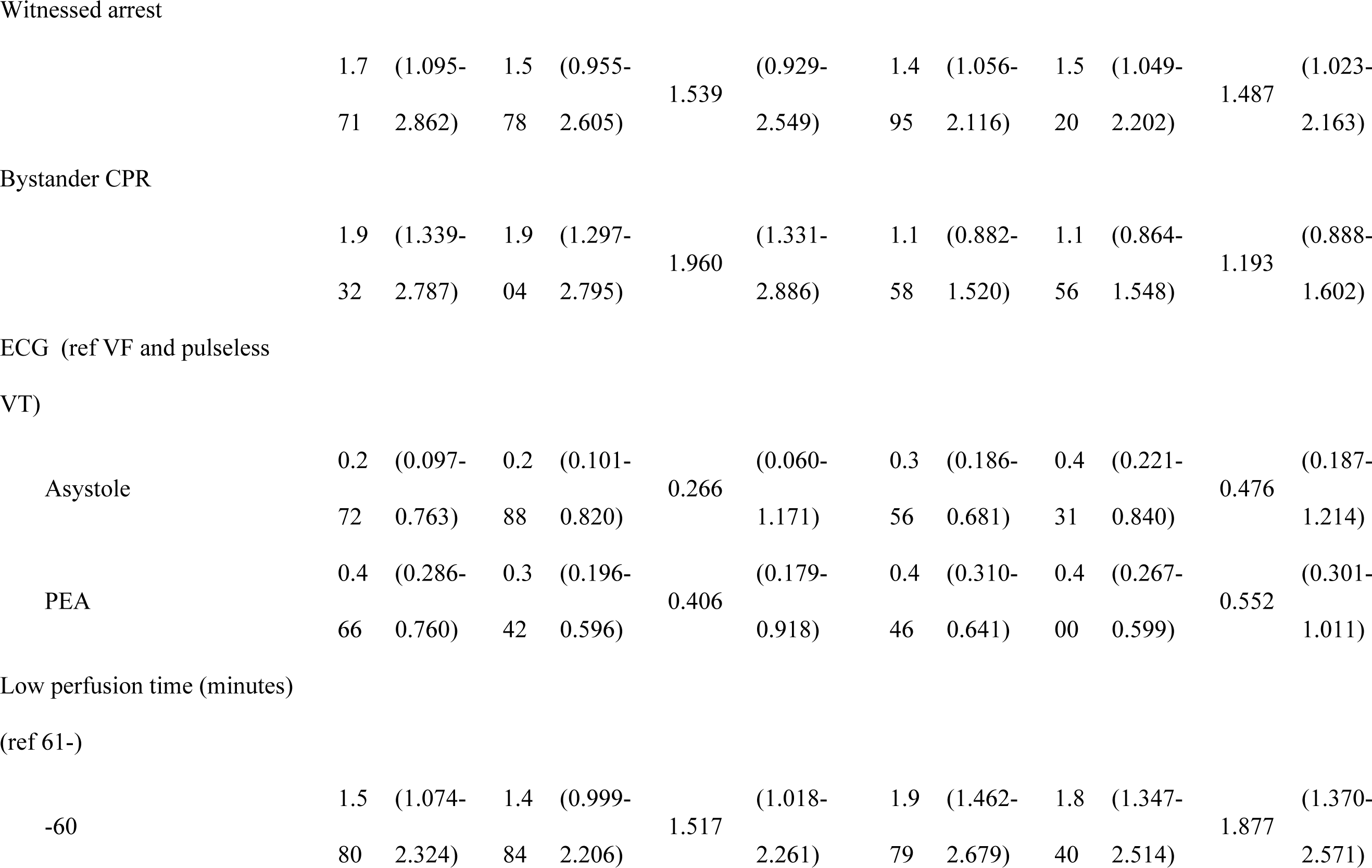
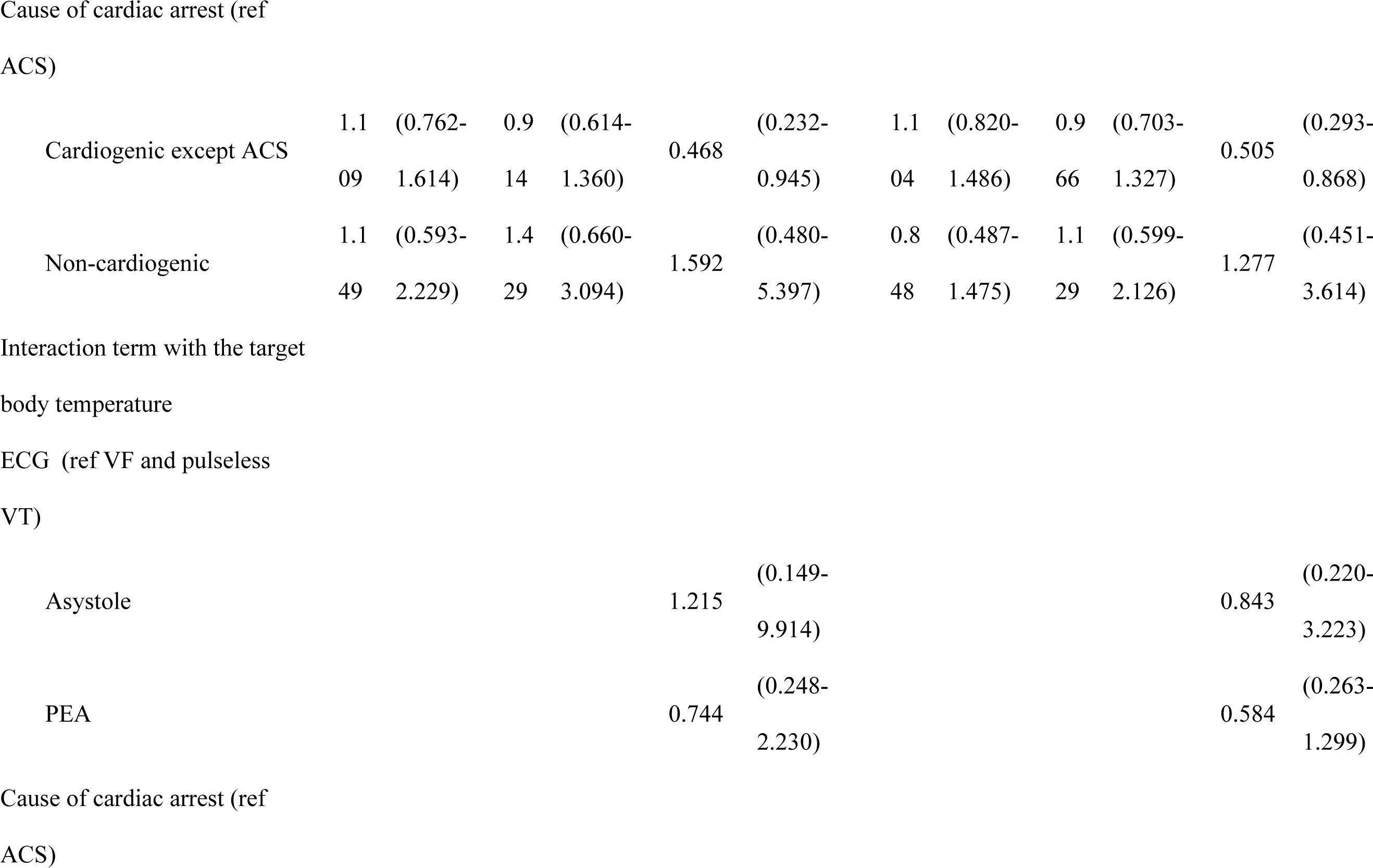
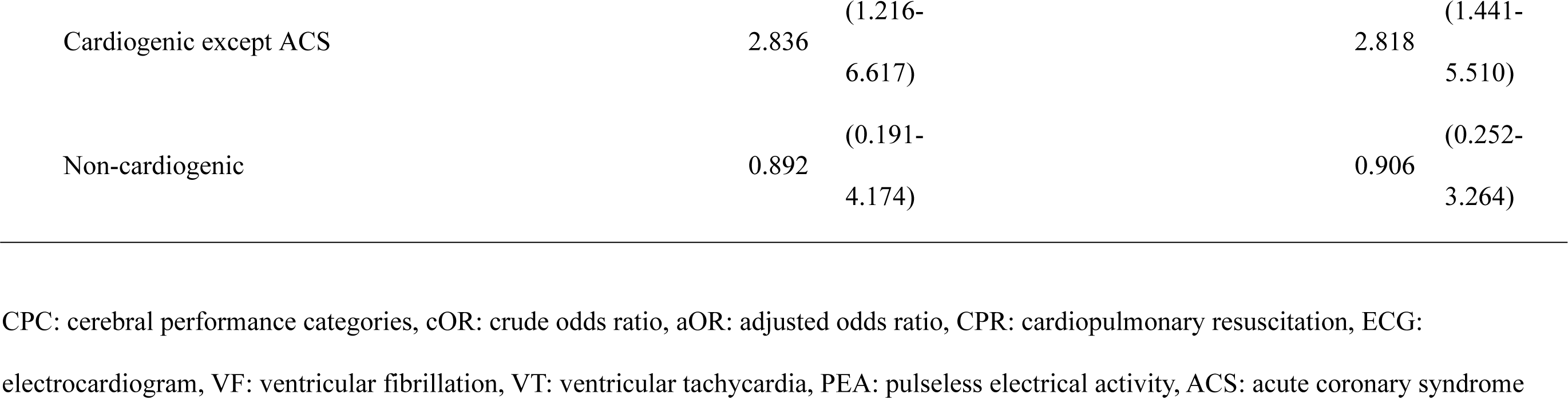
The odds ratio of factors which are likely to influence patient prognoses (ALL)

Of the interaction terms between target temperature, ECG, and causative disease, the only significant interaction with target temperature was identified as non-ACS cardiogenic cases for ACS in causative disease for good neurological outcome (aOR [95% CI] 2.8 [1.2– 6.6]) and for survival (aOR [95% CI] 2.8 [1.4–5.5]). (Table 2)

### Secondary Analysis

Based on the results of interaction, the 949 participants were stratified into two groups of 616 ACS and 269 non-ACS cardiogenic cases for the analysis; in the ACS cases, there was no intragroup difference in the proportion of patients with a favorable outcome. In the group of non-ACS cardiogenic cases at 35–36°C and 32–34°C, the favorable outcome rates were 10.3% and 24.2% and the survival rate was 19.8% and 50.3%, respectively (Table 3).

**Table 3.**
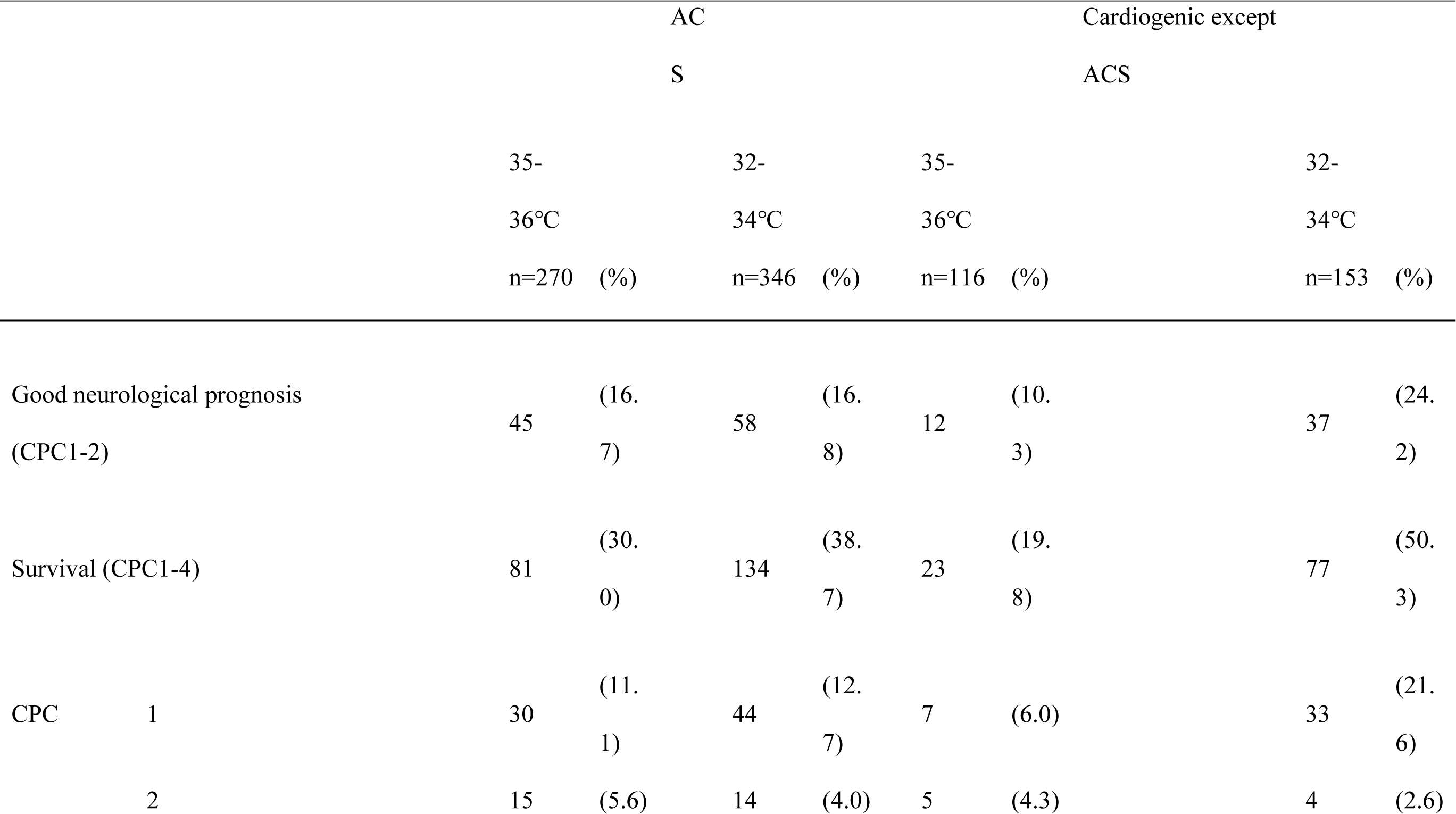
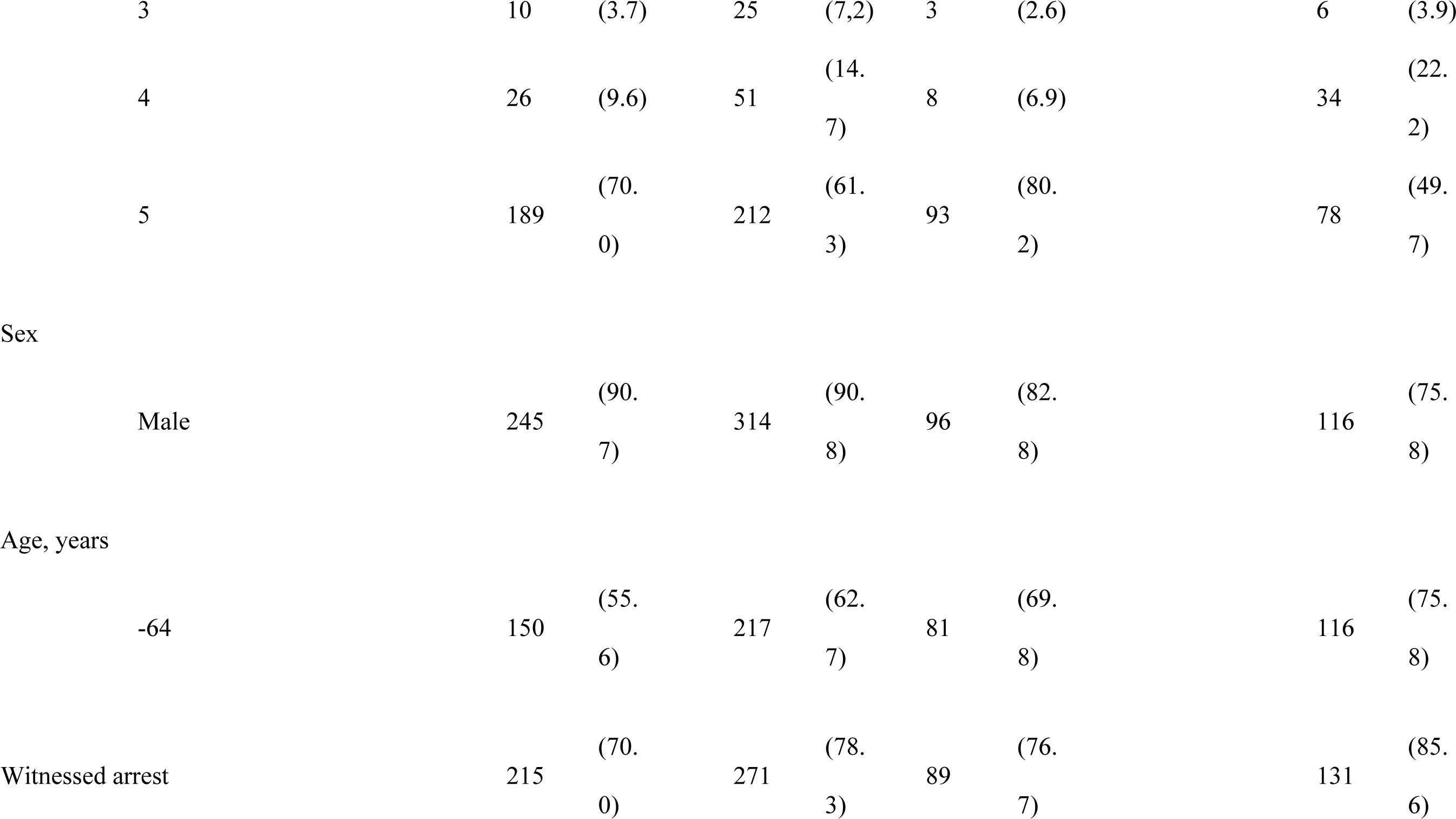
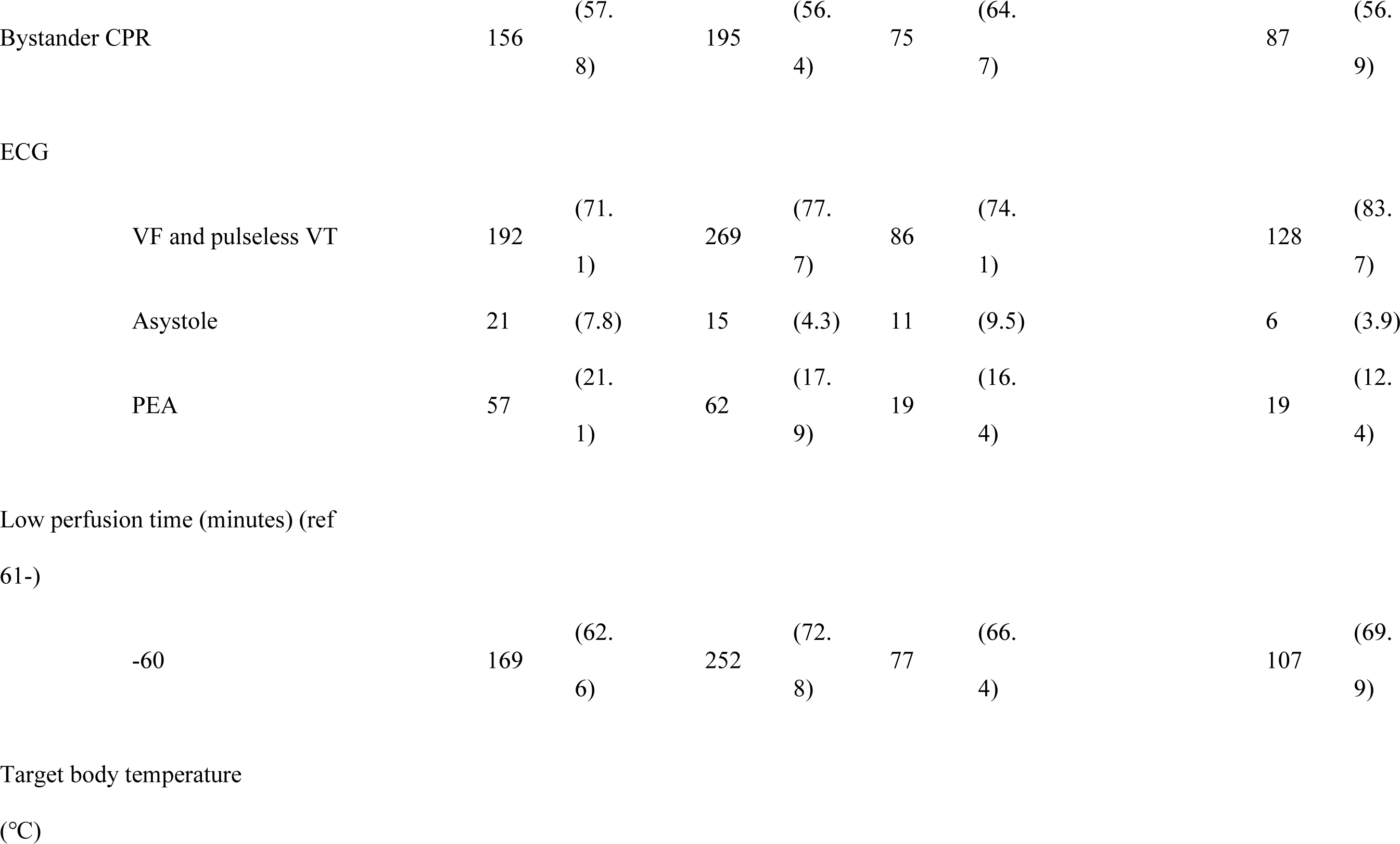
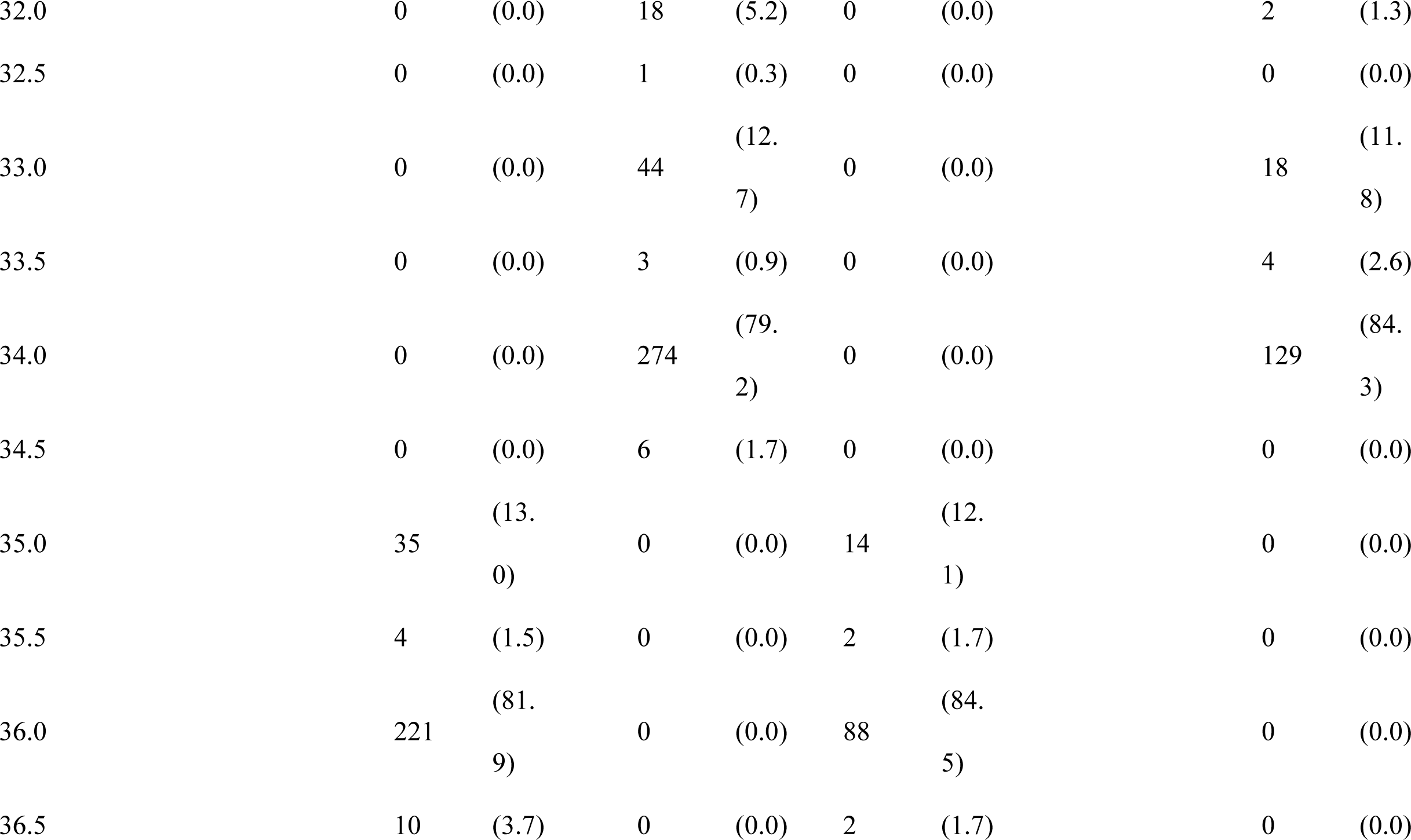
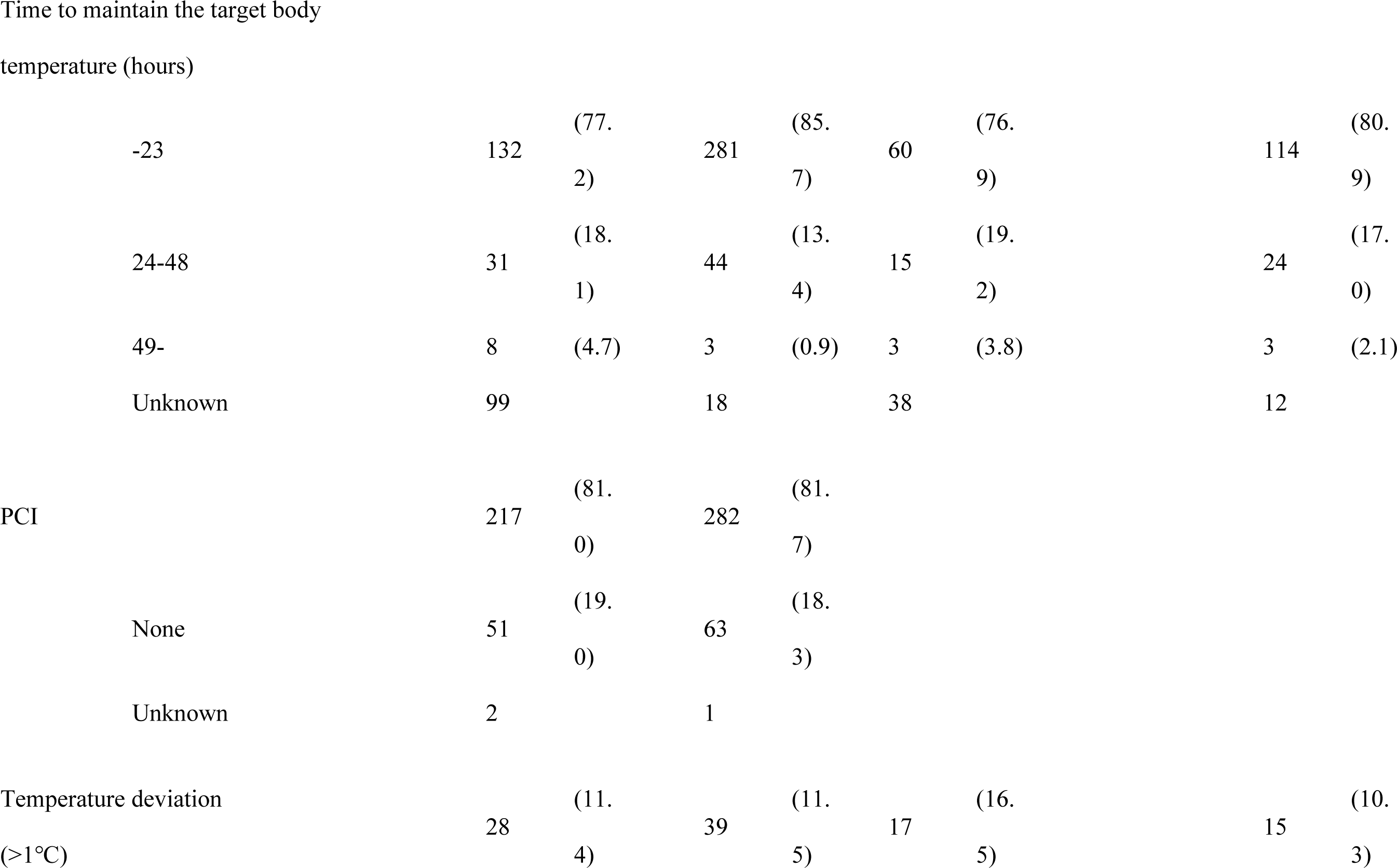
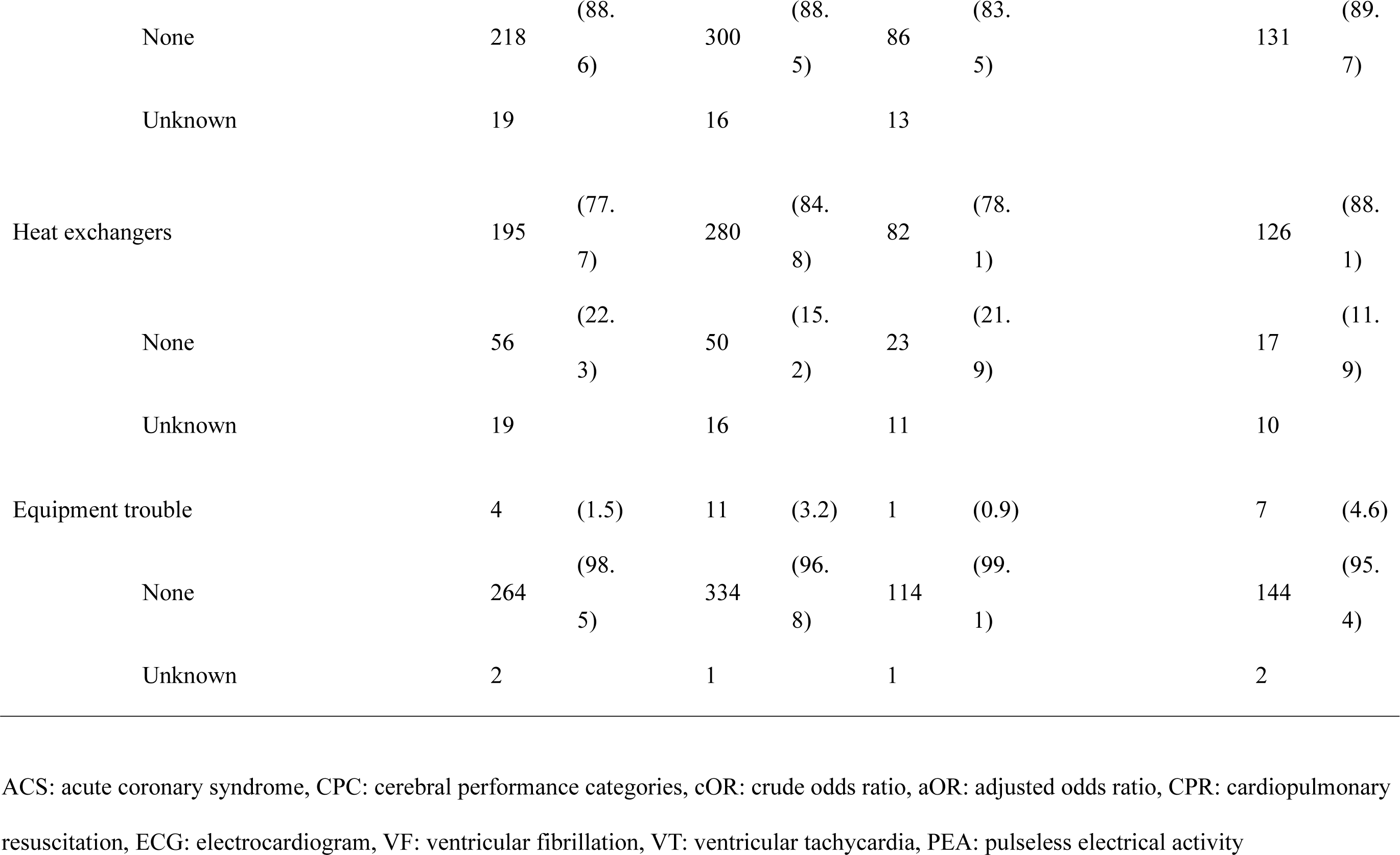
Description of participant characteristics in the ACS and cardiogenic shock (except ACS) groups.

|  |  | AC<br>S |  |  |  | Cardiogenic except<br>ACS |  |  |  |
| --- | --- | --- | --- | --- | --- | --- | --- | --- | --- |
|  |  | 35-36°C<br>n=270 (%) |  | 32-34°C<br>n=346 (%) |  | 35-36°C<br>n=116 (%) |  | 32-34°C<br>n=153 (%) |  |
| Good neurological prognosis<br>(CPC1-2) |  | 45 | (16.7) | 58 | (16.8) | 12 | (10.3) | 37 | (24.2) |
| Survival (CPC1-4) |  | 81 | (30.0) | 134 | (38.7) | 23 | (19.8) | 77 | (50.3) |
| CPC | 1 | 30 | (11.1) | 44 | (12.7) | 7 | (6.0) | 33 | (21.6) |
|  | 2 | 15 | (5.6) | 14 | (4.0) | 5 | (4.3) | 4 | (2.6) |
|  | 3 | 10 | (3.7) | 25 | (7,2) | 3 | (2.6) | 6 | (3.9) |
|  | 4 | 26 | (9.6) | 51 | (14.7) | 8 | (6.9) | 34 | (22.2) |
|  | 5 | 189 | (70.0) | 212 | (61.3) | 93 | (80.2) | 78 | (49.7) |
| Sex |  |  |  |  |  |  |  |  |  |
|  | Male | 245 | (90.7) | 314 | (90.8) | 96 | (82.8) | 116 | (75.8) |
| Age, years |  |  |  |  |  |  |  |  |  |
|  | -64 | 150 | (55.6) | 217 | (62.7) | 81 | (69.8) | 116 | (75.8) |
| Witnessed arrest |  | 215 | (70.0) | 271 | (78.3) | 89 | (76.7) | 131 | (85.6) |

|  |  |  |  |  |  |  |  |  |
| --- | --- | --- | --- | --- | --- | --- | --- | --- |
| Bystander CPR | 156 | (57.8) | 195 | (56.4) | 75 | (64.7) | 87 | (56.9) |
| ECG |  |  |  |  |  |  |  |  |
| VF and pulseless VT | 192 | (71.1) | 269 | (77.7) | 86 | (74.1) | 128 | (83.7) |
| Asystole | 21 | (7.8) | 15 | (4.3) | 11 | (9.5) | 6 | (3.9) |
| PEA | 57 | (21.1) | 62 | (17.9) | 19 | (16.4) | 19 | (12.4) |
| Low perfusion time (minutes) (ref 61-) |  |  |  |  |  |  |  |  |
| -60 | 169 | (62.6) | 252 | (72.8) | 77 | (66.4) | 107 | (69.9) |
| Target body temperature (°C) |  |  |  |  |  |  |  |  |
| 32.0 | 0 | (0.0) | 18 | (5.2) | 0 | (0.0) | 2 | (1.3) |
| 32.5 | 0 | (0.0) | 1 | (0.3) | 0 | (0.0) | 0 | (0.0) |
| 33.0 | 0 | (0.0) | 44 | (12.7) | 0 | (0.0) | 18 | (11.8) |
| 33.5 | 0 | (0.0) | 3 | (0.9) | 0 | (0.0) | 4 | (2.6) |
| 34.0 | 0 | (0.0) | 274 | (79.2) | 0 | (0.0) | 129 | (84.3) |
| 34.5 | 0 | (0.0) | 6 | (1.7) | 0 | (0.0) | 0 | (0.0) |
| 35.0 | 35 | (13.0) | 0 | (0.0) | 14 | (12.1) | 0 | (0.0) |
| 35.5 | 4 | (1.5) | 0 | (0.0) | 2 | (1.7) | 0 | (0.0) |
| 36.0 | 221 | (81.9) | 0 | (0.0) | 88 | (84.5) | 0 | (0.0) |
| 36.5 | 10 | (3.7) | 0 | (0.0) | 2 | (1.7) | 0 | (0.0) |

|  |  |  |  |  |  |  |  |  |
| --- | --- | --- | --- | --- | --- | --- | --- | --- |
| -23 | 132 | (77.<br>2) | 281 | (85.<br>7) | 60 | (76.<br>9) | 114 | (80.<br>9) |
| 24-48 | 31 | (18.<br>1) | 44 | (13.<br>4) | 15 | (19.<br>2) | 24 | (17.<br>0) |
| 49- | 8 | (4.7) | 3 | (0.9) | 3 | (3.8) | 3 | (2.1) |
| Unknown | 99 |  | 18 |  | 38 |  | 12 |  |
| PCI | 217 | (81.<br>0) | 282 | (81.<br>7) |  |  |  |  |
| None | 51 | (19.<br>0) | 63 | (18.<br>3) |  |  |  |  |
| Unknown | 2 |  | 1 |  |  |  |  |  |
| Temperature deviation<br>(>1°C) | 28 | (11.<br>4) | 39 | (11.<br>5) | 17 | (16.<br>5) | 15 | (10.<br>3) |
|  | None | 218 | (88.6) | 300 | (88.5) | 86 | (83.5) | 131 | (89.7) |
|  | Unknown | 19 |  | 16 |  | 13 |  |  |  |
| Heat exchangers |  | 195 | (77.7) | 280 | (84.8) | 82 | (78.1) | 126 | (88.1) |
|  | None | 56 | (22.3) | 50 | (15.2) | 23 | (21.9) | 17 | (11.9) |
|  | Unknown | 19 |  | 16 |  | 11 |  | 10 |  |
| Equipment trouble |  | 4 | (1.5) | 11 | (3.2) | 1 | (0.9) | 7 | (4.6) |
|  | None | 264 | (98.5) | 334 | (96.8) | 114 | (99.1) | 144 | (95.4) |
|  | Unknown | 2 |  | 1 |  | 1 |  | 2 |  |

ACS: acute coronary syndrome, CPC: cerebral performance categories, cOR: crude odds ratio, aOR: adjusted odds ratio, CPR: cardiopulmonary
resuscitation, ECG: electrocardiogram, VF: ventricular fibrillation, VT: ventricular tachycardia, PEA: pulseless electrical activity

Multivariate analysis in the ACS group showed that temperature control was not associated with either neurological outcome or survival (Table 4). However, in the group with non-ACS cardiogenic cases, the aOR for good neurological outcome and survival at 32–34°C versus 35–36°C was 2.9 (95% CI: 1.4–6.0) and 3.9 (95% CI: 2.2–6.9), respectively (Table 5).

**Table 4.** Odds ratio of factors that are likely to influence patient prognoses (ACS).

|  | Good neurological prognosis<br>(CPC1-2) |  |  |  | Survival (CPC1-4) |  |  |  |
| --- | --- | --- | --- | --- | --- | --- | --- | --- |
|  | cOR |  | aOR |  | cOR |  | aOR |  |
| Target body temperature (ref 35-36°C) |  |  |  |  |  |  |  |  |
| 32-34°C | 1.00<br>7 | (0.657-1.543) | 0.907 | (0.581-1.415) | 1.00<br>7 | (0.657-1.543) | 1.33<br>8 | (0.940-1.905) |
| Sex (ref female) |  |  |  |  |  |  |  |  |
| Male | 0.58<br>2 | (0.305-1.108) | 0.453 | (0.228-0.899) | 0.58<br>2 | (0.305-1.108) | 0.51<br>5 | (0.286-0.925) |
| Age (ref 65-) |  |  |  |  |  |  |  |  |
| -64 | 1.46<br>4 | (0.816-2.503) | 1.293 | (0.809-2.066) | 1.45<br>4 | (0.937-2.288) | 1.16<br>7 | (0.814-1.675) |
| Witnessed arrest |  |  |  |  |  |  |  |  |
|  | 1.42<br>9 | (0.816-2.503) | 1.350 | (0.749-2.435) | 1.42<br>9 | (0.816-2.503) | 1.44<br>0 | (0.922-2.251) |
| Bystander CPR |  |  |  |  |  |  |  |  |
|  | 1.94<br>8 | (1.235-3.072) | 1.967 | (1.223-3.162) | 1.94<br>8 | (1.235-3.072) | 1.29<br>1 | (0.902-1.847) |
| ECG (ref VF and pulseless VT) |  |  |  |  |  |  |  |  |
| Asystole | 0.11<br>5 | (1.074-2.324) | 0.116 | (0.016-0.870) | 0.11<br>5 | (0.015-0.847) | 0.50<br>2 | (0.219-1.146) |
| PEA | 0.36<br>8 | (0.185-0.731) | 0.317 | (0.155-0.645) | 0.36<br>8 | (0.185-0.731) | 0.34<br>5 | (0.207-0.573) |
| Low perfusion time (minutes) (ref<br>61-) |  |  |  |  |  |  |  |  |
| -60 | 1.58<br>0 | (1.704-2.324) | 1.608 | (0.968-2.670) | 1.58<br>0 | (1.074-2.324) | 2.04<br>8 | (1.381-3.037) |

ACS: acute coronary syndrome, CPC: cerebral performance categories, cOR: crude odds ratio, aOR: adjusted odds ratio, CPR: cardiopulmonary
resuscitation, ECG: electrocardiogram, VF: ventricular fibrillation, VT: ventricular tachycardia, PEA: pulseless electrical activity

**Table 5.** Odds ratio of factors that are likely to influence patient prognoses (cardiogenic shock, except ACS).

|  | Good neurological prognosis<br>(CPC1-2) |  |  |  | Survival (CPC1-4) |  |  |  |
| --- | --- | --- | --- | --- | --- | --- | --- | --- |
|  | cOR |  | aOR |  | cOR |  | aOR |  |
| Target body temperature (ref 35-36°C) |  |  |  |  |  |  |  |  |
| 32-34°C | 2.76<br>4 | (1.369-5.583) | 2.870 | (1.362-6.049) | 4.09<br>7 | (2.350-7.141) | 3.86<br>7 | (2.161-6.919) |
| Sex (ref female) |  |  |  |  |  |  |  |  |
| male | 0.53<br>5 | (0.267-1.071) | 0.558 | (0.264-1.179) | 0.84<br>3 | (0.463-1.534) | 0.92<br>3 | (0.479-1.776) |
| Age (ref 65-) |  |  |  |  |  |  |  |  |
| -64 | 1.32<br>5 | (0.637-2.759) | 1.194 | (0.537-2.655) | 1.49<br>0 | (0.837-2.653) | 1.35<br>8 | (0.717-2.569) |
| Witnessed arrest |  |  |  |  |  |  |  |  |
|  | 1.42<br>9 | (0.816-2.503) | 2.829 | (0.806-9.932) | 1.42<br>9 | (0.816-2.503) | 1.87<br>2 | (0.858-4.082) |
| Bystander CPR |  |  |  |  |  |  |  |  |
|  | 2.67<br>4 | (1.299-5.506) | 2.890 | (1.327-6.925) | 1.05<br>3 | (0.635-1.747) | 1.16<br>2 | (0.652-2.070) |
| ECG (ref VF and pulseless VT) |  |  |  |  |  |  |  |  |
| Asystole | 0.51<br>5 | (0.114-2.337) | 0.665 | (0.138-3.199) | 0.30<br>1 | (0.084-1.078) | 0.42<br>0 | (0.109-1.615) |
| PEA | 0.33<br>1 | (0.097-1.127) | 0.283 | (0.080-1.008) | 0.37<br>5 | (0.164-3.714) | 0.39<br>7 | (0.164-0.957) |
| Low perfusion time (minutes) (ref<br>61-) |  |  |  |  |  |  |  |  |
| -60 | 1.53<br>1 | (0.754-3.111) | 1.487 | (0.698-3.169) | 2.10<br>7 | (1.196-3.714) | 2.04<br>9 | (1.114-3.767) |

ACS: acute coronary syndrome, CPC: cerebral performance categories, cOR: crude odds ratio, aOR: adjusted odds ratio, CPR: cardiopulmonary
resuscitation, ECG: electrocardiogram, VF: ventricular fibrillation, VT: ventricular tachycardia, PEA: pulseless electrical activity

## Conclusions

In the main analysis, although the target body temperature of 32–34°C (hypothermic management) was not a significant factor for better neurological outcome as compared to management at 35–36°C (normothermic control), this was a significant outcome-related factor for survival. On the other hand, when disease was considered, cardiogenicity due to ACS was not a significant factor for either neurological outcome or survival whereas cardiogenicity other than ACS (arrhythmia, myopathy, myocarditis, and other cardiac causes) was a significant factor for both neurological outcome and survival.

Traditionally, hypothermic management has been aimed at a cerebroprotective effect. When cerebral blood flow is disrupted by cardiac arrest, anaerobic glycolysis occurs, inducing structural changes in the Na+/K+ATPase and Ca^2+^-ATPase and triggering glutamate release into the extracellular space, which results in a sustained intracellular Ca^2+^ influx. The increase in intracellular Ca^2+^ concentration leads to lipid peroxide and free radical production, which results in cell death.^16^ Hypothermic management likely confers a brain protective effect by decreasing brain metabolism, and suppressing oxygen consumption and glutamate release.^17^ However, the cerebroprotective effect alone is insufficient to explain why hypothermic management constitutes a significant prognostic factor for survival. On the other hand, myocardial injury progresses due to a decrease in the myocardial ATP content, secondary to the progression of anaerobic glycolysis due to the disruption of coronary artery blood flow. This myocardial injury has both reversible and irreversible components, wherein the former comprises stunned myocardium and the latter includes cell death. To improve the outcome after cardiac arrest, it is important to improve the rate of recovery of cardiac function from the state of myocardial stunning.^18^ Experimental studies examining the effects of ischemia time and ischemia temperature on the rate of recovery of cardiac function after ischemia and reperfusion, have shown that the rate of recovery of cardiac function and residual ATP content are not reduced with prolonged ischemia time at low temperature, whereas at room temperature, both the rate of cardiac function recovery and residual ATP content are significantly reduced by prolonged ischemia time.^19^ This myocardial protective effect of hypothermic control may be related to the fact that hypothermic control was a significant prognostic factor for survival in the present study.

In ACS, cell death is often accompanied by impaired cell membrane damage and deviations in intracellular enzymatic activity. In particular, the ACS patients in this study were already in a state of cardiac arrest, and even if urgent PCI was performed, many of them had myocardial cell death; moreover, mechanical complications such as acute mitral regurgitation due to papillary muscle tear and ventricular septal defect may have been associated with the outcomes and the hypothermic management of myocardial stunting. The impact of improved recovery of cardiac function from the condition may have been limited. However, in myopathy and myocarditis, if the patient survives the acute stage, the disease will spontaneously alleviate and likely ensure a favorable outcome. This is because in myopathy and myocarditis, hypoxia due to pump failure worsens and leads to cardiac arrest, which results in fewer myocardial cells experiencing cell death and more recovery of cardiac function from the state of myocardial stunting. Therefore, in cardiomyopathies other than ACS, hypothermic management may have been a significant factor for both neurological outcome and survival compared to normothermic management.

The results of previous studies on TTM after cardiac arrest are not uniform, and the results of this study may appear to contradict the findings of some previous studies. First, the uniqueness of this study is that the analysis was performed with stable circulatory and temperature management by ECMO, and taking into account the underlying disease; by using a heat exchanger in the ECMO circuit at 80%, the deviation of more than 1°C was limited to approximately 10%; therefore, we infer that stable temperature control was achieved. On the other hand, as hypothermic management suppresses circulatory dynamics, many patients in previous studies that did not use ECMO may have suffered cardiac arrest again before recovery from myocardial stunting. The TTM-1 Study, TTM-2 Study, and part of the JAAM-OHCA Registry reported that hypothermia management was not effective,^6, 7, 10^ while the HYPERION trial and another part of the JAAM-OHCA Registry reported that hypothermia management was effective.^9, 10^ The TTM-1 trial included patients with cardiac arrest of presumed cardiac cause, the TTM-2 trial included patients with cardiac arrest of presumed or unknown cause, regardless of initial rhythm,^7, 8^ and the HYPERION trial included patients resuscitated from non-shocking rhythm cardiac arrest from any cause and does not consider whether the patient has ACS.^9^ The OHCA Registry report analyzed patients according to whether ECMO was performed and severity of illness, and did not consider causative illness.^10^ Therefore, when the discrepancies in these series of reports on the effectiveness of hypothermia management are examined in light of the results of this study, it is presumed that the composition of causative diseases in each report was different, which may have affected the effectiveness of hypothermia management. Regarding the JAAM-OHCA Registry study that focused on patients who underwent TTM with ECMO, it was reported that hypothermic management was not necessary. However, as in other previous studies, whether the causative disease was ACS was not considered. Although this report does not exclude ROSC cases prior to the introduction of ECMO, which was excluded in this study, and as a simple prognostic comparison cannot be made, the neurological outcome (CPC1-2) at 30 days for both the 32–34°C and 35–36°C groups was 15% and the 30-day survival rate was 35%, which is considered to be the same level as the survival and good outcome rates in the ACS of this study.^11^

There are three limitations of this study. The first is that because the criteria for ECMO implementation and the method of temperature management are left to the institutional standards, the inclusion and distribution of patients may be arbitrary in such a way that hypothermic management was implemented for patients who have witnessed bystander CPR, and are expected to have a good outcome. Although this study addresses the variables of witnessed arrest, bystander CPR, age, and low perfusion time, there may be other factors that influence the inclusion of hypothermia management.

Second, there is no information on the method of temperature management, such as the time to maintain the target body temperature and the site of temperature measurement. However, we confirmed that more than 80% of the target body temperatures were 34.0°C for hypothermic management and 36.0°C for normothermic management, and that there was no significant difference in maintenance time, use of heat exchangers, or temperature deviation rates between the two groups of hypothermia and normothermia managements. Thus, we can assume that differences in measurement and control methods are small.

Third, the degree of myocardial necrosis was not examined quantitatively. The present study assumes that the degree of myocardial necrosis in ACS is high, and the classification of the causative disease can be considered to outline the degree of myocardial necrosis. However, the degree of myocardial necrosis may differ even when inpatients are classified as having the same ACS, and myocardial necrosis may be advanced in other cardiogenic conditions besides ACS that lead to cardiac arrest. Quantitative studies would be possible by measuring myocardial deviation enzymes and other parameters.

As described above, although the influence of limitation cannot be completely eliminated in the use of registry data, we judged that we were able to minimize its influence so that it did not detract from the usefulness of the results of this study. However, prospective validation is needed with a clearly defined plan that includes the criteria for the introduction of ECMO, methods of temperature control, and validation of the extent of myocardial necrosis.

In temperature management using ECMO after ECPR induction, target body temperature of 32–34°C (hypothermic management) was a significant prognostic determinant for survival as compared to that of 35–36°C (normothermic management). In addition, neither management was a significant factor for neurological outcome and survival in cardiogenicity due to ACS, although both were significant factors for good neurological outcome and survival in cardiogenicity other than ACS (arrhythmia, myopathy, myocarditis, and other cardiac causes). Myocardial protection by hypothermic management may have an effect.

## Data Availability

The data that support the findings of this study are available from the corresponding author upon reasonable request.

## Acknowledgments

Hirotaka Sawano, M.D., Ph.D. (Osaka Saiseikai Senri Hospital), Yuko Egawa, M.D., Shunichi Kato, M.D. (Saitama Red Cross Hospital), Naofumi Bunya, M.D., Takehiko Kasai, M.D. (Sapporo Medical University), Shinichi Ijuin, M.D., Shinichi Nakayama, M.D., Ph.D. (Hyogo Emergency Medical Center), Seiya Kanou, M.D. (Teikyo University Hospital), Toru Takiguchi, M.D.(Nippon Medical School), Hiroaki Takada, M.D., Kazushige Inoue, M.D. (National Hospital Organization Disaster Medical Center), Ichiro Takeuchi, M.D., Ph.D., Hiroshi Honzawa, M.D. (Yokohama City University Medical Center), Makoto Kobayashi, M.D., Ph.D., Tomohiro Hamagami, M.D. (Toyooka Public Hospital), Wataru Takayama, M.D., Yasuhiro Otomo, M.D., Ph.D. (Tokyo Medical and Dental University Hospital of Medicine), Kunihiko Maekawa, M.D. (Hokkaido University Hospital), Takafumi Shimizu, M.D., Satoshi Nara, M.D. (Teine Keijinkai Hospital), Michitaka Nasu, M.D., Kuniko Takahashi, M.D. (Urasoe General Hospital), Yoshihiro Hagiwara, M.D., M.P.H. (Imperial Foundation Saiseikai, Utsunomiya Hospital), Shigeki Kushimoto, M.D., Ph.D. (Tohoku University Graduate School of Medicine), Reo Fukuda, M.D. (Nippon Medical School Tama Nagayama Hospital), Takayuki Ogura, M.D., Ph.D. (Japan Red Cross Maebashi Hospital), Shin-ichiro Shiraishi, M.D. (Aizu Central Hospital), Ryosuke Zushi, M.D. (Osaka Mishima Emergency Critical Care Center), Norio Otani, M.D. (St. Luke’s International Hospital), Kazuhiro Watanabe, M.D. (Nihon University Hospital), Takuo Nakagami, M.D. (Omihachiman Community Medical Center), Tomohisa Shoko, M.D., Ph.D. (Tokyo Women’s Medical University Medical Center East), Nobuya Kitamura, M.D., Ph.D. (Kimitsu Chuo Hospital), Takayuki Otani, M.D. (Hiroshima City Hiroshima Citizens Hospital), Yoshinori Matsuoka, M.D., Ph.D. (Kobe City Medical Center General Hospital), Makoto Aoki, M.D., Ph.D. (Gunma University Graduate School of Medicine), Masaaki Sakuraya, M.D., M.P.H. (JA Hiroshima General Hospital Hiroshima), Hideki Arimoto, M.D. (Osaka City General Hospital), Koichiro Homma, M.D., Ph.D. (Keio University School of Medicine), Hiromichi Naito, M.D., Ph.D. (Okayama University Hospital), Shunichiro Nakao, M.D., Ph.D. (Osaka University Graduate School of Medicine), Tomoya Okazaki, M.D., Ph.D. (Kagawa University Hospital), Yoshio Tahara, M.D., Ph.D. (National Cerebral and Cardiovascular Center), Hiroshi Okamoto, M.D, M.P.H. (St. Luke’s International Hospital), Jun Kunikata, M.D., Ph.D., Hideto Yokoi, M.D., Ph.D. (Kagawa University Hospital).

## Sources of Funding

This work was supported by the Japan Society for the Promotion of Science KAKENHI to JK(JP19K18365) and YK (JP19K09419). The funding organization did not play any role in the study; the views expressed in this paper do not reflect the views of the Ministry.

## Disclosures

None

## Notes

### Competing Interest Statement

The authors have declared no competing interest.

